# Breathing zone pollutant levels are associated with asthma exacerbations in high-risk children

**DOI:** 10.1101/2023.09.22.23295971

**Authors:** Camille M. Moore, Jonathan Thornburg, Elizabeth A. Secor, Katharine L. Hamlington, Allison M. Schiltz, Kristy L. Freeman, Jamie L. Everman, Tasha E. Fingerlin, Andrew H. Liu, Max A. Seibold

## Abstract

**Background:** Indoor and outdoor air pollution levels are associated with poor asthma outcomes in children. However, few studies have evaluated whether breathing zone pollutant levels associate with asthma outcomes.

**Objective:** Determine breathing zone exposure levels of NO_2_, O_3_, total PM_10_ and PM_10_ constituents among children with exacerbation-prone asthma, and examine correspondence with in-home and community measurements and associations with outcomes.

**Methods:** We assessed children’s personal breathing zone exposures using wearable monitors. Personal exposures were compared to in-home and community measurements and tested for association with lung function, asthma control, and asthma exacerbations.

**Results:** 81 children completed 219 monitoring sessions. Correlations between personal and community levels of PM_10_, NO_2_, and O_3_ were poor, whereas personal PM_10_ and NO_2_ levels correlated with in-home measurements. However, in-home monitoring underdetected brown carbon (Personal:79%, Home:36.8%) and ETS (Personal:83.7%, Home:4.1%) personal exposures, and detected black carbon in participants without these personal exposures (Personal: 26.5%, Home: 96%). Personal exposures were not associated with lung function or asthma control. Children experiencing an asthma exacerbation within 60 days of personal exposure monitoring had 1.98, 2.21 and 2.04 times higher brown carbon (p<0.001), ETS (p=0.007), and endotoxin (p=0.012), respectively. These outcomes were not associated with community or in-home exposure levels.

**Conclusions:** Monitoring pollutant levels in the breathing zone is essential to understand how exposures influence asthma outcomes, as agreement between personal and in-home monitors is limited. Inhaled exposure to PM_10_ constituents modifies asthma exacerbation risk, suggesting efforts to limit these exposures among high-risk children may decrease their asthma burden.

**CLINICAL IMPLICATIONS:** In-home and community monitoring of environmental pollutants may underestimate personal exposures. Levels of inhaled exposure to PM_10_ constituents appear to strongly influence asthma exacerbation risk. Therefore, efforts should be made to mitigate these exposures.

**CAPSULE SUMMARY:** Leveraging wearable, breathing-zone monitors, we show exposures to inhaled pollutants are poorly proxied by in-home and community monitors, among children with exacerbation-prone asthma. Inhaled exposure to multiple PM_10_ constituents is associated with asthma exacerbation risk.

## INTRODUCTION

Asthma is one of the most prevalent diseases of childhood, impacting an estimated 6 million children in the United States.^1^ For many, asthma can be effectively controlled using standard therapies; however, it is estimated that up to 18% of children with asthma have more severe disease, characterized by frequent exacerbations that require emergency department visits, courses of systemic corticosteroids, and/or hospitalization.^2–4^ These exacerbation-prone (EP) children experience the majority of the patient burden and economic costs associated with childhood asthma.^5^ In addition, asthma exacerbations are associated with progressive loss of lung function, which puts children with EP-asthma at increased risk for chronic lung disease in adulthood.^6–8^

While viruses, particularly human rhinoviruses A and C, are implicated in the majority of childhood asthma exacerbations^9, 10^, non-viral exacerbations may be triggered by air pollution.^11^ In addition, air pollution may impair mucosal barrier function^12, 13^ and phagocyte airway clearance^14^, as well as alter adaptive immune responses^15, 16^, suggesting that air pollution could indirectly influence risk for exacerbations by increasing susceptibility to and harm from viral infections^17–19^.

Several epidemiological studies found that peaks in emergency department and hospital admission for asthma coincide with seasonal fluctuations in ambient air pollution and that increased levels of ambient pollutants are associated with exacerbation.^20, 21^ However, results have been mixed in terms of which pollutants are associated with increased risk.^22–29^ Studies of the impact of ambient pollution on asthma symptoms and lung function have also been inconsistent, with some studies finding air pollution results in poor asthma outcomes,^30–32^ others finding no impact,^33, 34^ and others finding associations only for specific subgroups of children.^35, 36^

One explanation for these conflicting results is exposure misclassification or measurement error due to using community ambient air quality monitoring sites to assess an individual’s exposure level.^37, 38^ Children spend a majority of their time indoors^39^, and there can be large differences between indoor and outdoor levels of air pollutants.^40–44^ Therefore, there has been a growing interest in assessing the impact of indoor air pollution and allergen levels on asthma outcomes.^32, 33, 45–50^ However, studies employing indoor stationary monitors placed in a single location (usually a child’s bedroom) suffer from many of the same problems as using outdoor monitors,^51–53^ as children spend a significant portion of their day away from the home in school and physical activity can generate a “personal cloud” of increased particulate matter (PM) exposure that may not be captured by a stationary monitor.^43, 54^ Inaccurate exposure measurements can bias results of analyses and mask associations between environmental exposures and asthma outcomes.^37^

In this study, we investigate the role of personal environmental exposures in asthma outcomes in an urban cohort of children with EP asthma. Person-level environmental exposures, including ozone (O_3_), nitrogen dioxide (NO_2_), PM_10_ (particles less than 10 microns in diameter), and several PM_10_ constituents, including brown carbon (BrC), black carbon (BC), environmental tobacco smoke (ETS), endotoxin and 𝛽-glucan, were collected using wearable monitors to accurately characterize individual children’s breathing zone exposure levels. We describe factors associated with personal exposures, assess the agreement between personal exposure measurements and community outdoor and in-home measures, and evaluate the relationship between personal exposures and asthma outcomes, including lung function, asthma severity, and exacerbation.

## METHODS

### Cohort

ENIGMA was an observational study of children with EP asthma conducted from June 2018 to September 2022 in the Denver metro area in Colorado. We recruited 100 children 8 to 16 years old with clinician-diagnosed asthma. Initially, eligible children were required to have had at least one asthma exacerbation in the prior 12 months; however, eligibility criteria were modified in December 2020 to include children with at least one exacerbation in the prior 18 months to aid in recruitment during the COVID-19 pandemic. An asthma exacerbation was defined as an asthma-related unscheduled visit to an emergency department, clinic, or urgent care facility; overnight hospitalization; or course of systemic corticosteroids. Individuals were excluded if they were self-reported active smokers, homeschooled, or had conditions that would interfere with the safety or performance of the study. Initially, our study design was longitudinal seasonal assessments of environmental exposures which were immediately followed by a clinical assessment of lung function and asthma status. Due to the COVID-19 pandemic, we switched to a cross-sectional design with modifications to the exposure and clinical assessments in February 2021. The Colorado Multiple Institutional Review Board approved the protocol for the study. The participant and at least one legal guardian provided informed written consent and, if age-appropriate, assent.

### Environmental Exposure Assessments

Personal exposure monitoring was performed for approximately 72 hours prior to all scheduled study visits performed through May 2022. RTI MicroPEMs™ (Research Triangle Park, NC) collected filter and real-time PM_10_, temperature, humidity, and accelerometry measurements. Ogawa passive dosimeters (Ocala, FL) measured NO_2_ and O_3_ (summer only) exposure.^55^ Participants wore all devices in a belt bag placed diagonally across the chest such that the monitor inlet was within the breathing zone i.e., within 10 inches of the nose and mouth.^56, 57^ PM_10_ filters underwent gravimetric analysis^58^ and were then analyzed for BC, BrC, ETS, endotoxin and 𝛽-glucan (Supplement). Pre-pandemic participants (n=49) also had in-home environmental monitors placed in their bedroom concurrent with their first 72-hour personal monitoring session. Hourly air quality data (O3, NO2, and PM10) from the Denver Colorado Air Monitoring Program monitoring site were obtained from the Environmental Protection Agency’s Air Quality System API for years 2018 through 2022.

### Clinical Assessments

Clinical assessments included lung function measured by spirometry, asthma control, and questions on healthcare and medication usage in the previous 60 days (Supplement). Spirometry was performed per American Thoracic Society/European Respiratory Society standards.^59^ Asthma control was assessed with the validated Asthma Control Test (ACT)^60^ for participants ages 12 years and older (5-25 scale), and the Childhood ACT (cACT)^61, 62^ for participants ages 6-11 years (0-27 scale).

### Statistical Methods

We compared log_10_(personal exposures) between participants with and without asthma control (cACT or ACT ≥ 20 vs. <20), hospitalization in the 18 months prior to enrollment, and signs of exacerbation in the 60-days prior to their exposure assessment (systemic corticosteroid use or unscheduled health care visits for asthma), using censored regression models to account for observations below the limit of detection (LOD) with robust standard errors to account for repeated measures. As over 50% of BC measurements were below the LOD, we dichotomized BC levels into “Detected” vs. “Not Detected” and used generalized estimating equation (GEE) logistic regression models. Pre-bronchodilator spirometry and the percent change in spirometry measures after bronchodilation were evaluated for association with the log_10_(exposures) using GEE models. All models controlled for age, sex, race-ethnicity, season (except O_3_) and monitor wearing compliance, measured as the percent of awake time spent wearing the personal monitor. In addition, post-bronchodilation change models were adjusted for the corresponding baseline lung function measure. A Benjamini-Hochberg correction was applied to the p-values across the eight exposures to control the false discovery rate (FDR) at 5%. Full details are available in the Supplement.

## RESULTS

### ENIGMA cohort demographic and clinical characteristics

The ENIGMA study performed personal exposure monitoring on 81 children with EP asthma. Participants ranged from 8.2 to 16.7 years in age (median: 12 years, interquartile range (IQR): 10.7-14.4; Table 1) and 64% were male. The majority reported Hispanic ethnicity (56.8%); 13.6%, 14.8%, and 14.8% reported non-Hispanic Black, non-Hispanic White, and other race/ethnicities, respectively. Out of the 81 participants, 75 (92.6%) experienced 1 or more exacerbation in the 12 months prior to enrollment. In addition, 88.9% had one or more unscheduled healthcare or emergency department visits, 86.4% had been prescribed oral corticosteroids, and 24.7% required hospitalization related to their asthma in the 18 months prior to enrollment. During the course of the study, 29.6% of participants reported one or more unscheduled asthma-related health care visits and 25.9% reported one or more courses of systemic corticosteroids for asthma in the 60 days prior to a study visit.

**Table 1:** Participant characteristics (N=81). N(%) or median [IQR].

| Variable | N (%) or Median (IQR) |
| --- | --- |
| Age (years) | 12.02 [10.72, 14.37] |
| Sex: Female | 29 (35.8) |
| Race / Ethnicity: Hispanic | 46 (56.8) |
| Non-Hispanic Black | 11 (13.6) |
| Non-Hispanic Other | 12 (14.8) |
| Non-Hispanic White | 12 (14.8) |
| CDC BMI Percentile | 79.90 [54.20, 93.50] |
| Household Income | \$45,000 [\$15,000 - \$19,999, \$55,000 - \$59,999] |
| Parental Education Level: Grades 1-11 | 10 (12.3) |
| GED or 12th grade | 26 (32.1) |
| >12th grade | 45 (55.6) |
| Medication Use | 17 (21.0) |
|  | 23 (28.4) |
|  | 10 (12.3) |
|  | 15 (18.5) |
|  | 16 (19.8) |
| Baseline Asthma Control<br>(cACT or ACT < 20) | 57 (72.2) |
|  | 22 (27.8) |
| FEV1 pp pre (subject median) | 94.37 [85.79, 108.01] |
| FVC pp pre (subject median) | 106.58 [95.57, 118.54] |
| FEV1/FVC pp pre (subject median) | 90.02 [85.14, 95.18] |
| FEV1 % Change Post Bronchodilator (subject median) | 8.23 [5.14, 12.70] |
| FVC % Change Post Bronchodilator (subject median) | 1.25 [-0.21, 3.57] |
| FEV1/FVC % Change Post Bronchodilator (subject median) | 7.37 [3.45, 10.44] |
| Number of Visits: 1 | 26 (32.1) |
| 2 | 8 ( 9.9) |
| 3 | 15 (18.5) |
| 4 | 28 (34.6) |
| 5 | 4 ( 4.9) |

### ENIGMA environmental exposure assessment design

In the pre-pandemic phase of ENIGMA (June 2018 to February 2020), children underwent clinical characterization and biosampling study visits every 3 months for up to 12 months (Figure 1). 72-hours prior to these study visits, participants were directed to wear personal monitors collecting ambient PM_10_, NO_2_ and O_3_ (summer only). Additionally, particulates on the monitor filter were analyzed to determine BC, BrC, and ETS, and extracted to measure endotoxin, and 𝛽-glucan levels. However, visits were paused during the first year of the COVID-19 pandemic (March 2020-February 2021), after which the study continued with a cross-sectional design. Therefore, longitudinal exposure data is available on 55 participants and single timepoint data on 26 participants.

**Figure 1.**
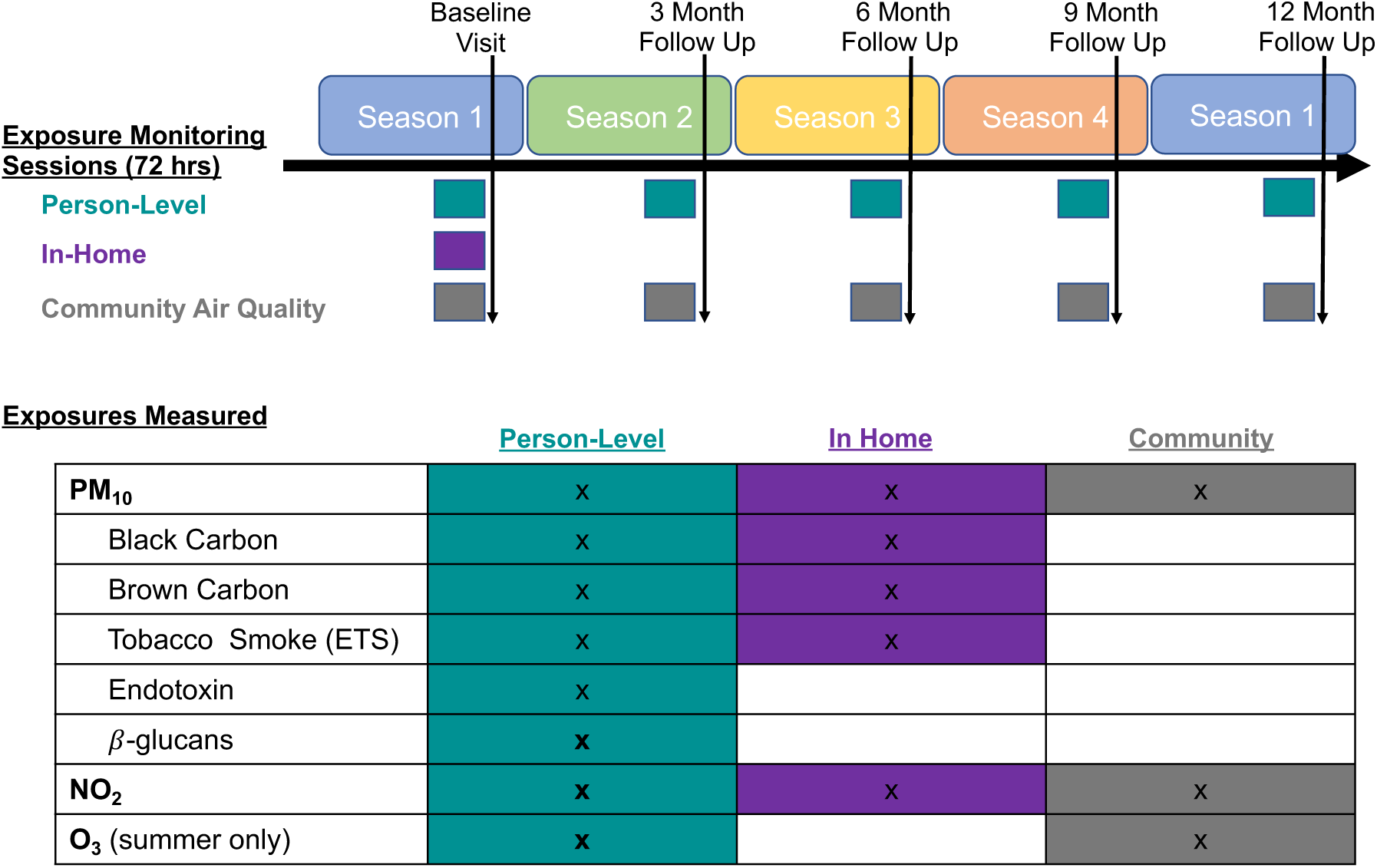
Overview of the ENIGMA study design.

For comparison to the personal monitor data, we obtained community air quality monitoring station data (PM_10_, NO_2_, O_3_), matched to the participants’ personal monitoring periods. Additionally, for most pre-pandemic participants (N=49), we also collected in-home exposure levels (PM_10_, NO_2_, BC, BrC, and ETS), measured from stationary monitors placed near and in the same room as the child’s bed, during the same time as the first personal monitoring session.

### Personal exposure to environmental pollutants is widespread and highly variable in children with EP-asthma

In total, 219 personal exposure monitoring sessions were conducted across the 81 participants, with a median of 3 monitoring sessions per participant, pre-pandemic. Participants had high wearing compliance, wearing their personal monitors for a median of 72.1% (IQR: 53.2%-88.4%) of their time awake.

99.5%, 98.5%, and 94.8% of the collected samples had detectable PM_10_, NO_2_, and O_3_, respectively (Figure 2A, Table 2). However, there was wide variation in exposure level for each, with PM_10_ ranging from <1 to 141.9 𝜇g/m^3^, NO_2_, from <2 to 99.1 parts per billion (ppb), and O_3_ from <1.5 to 23.3 ppb (Figure 2B).

**Figure 2.**
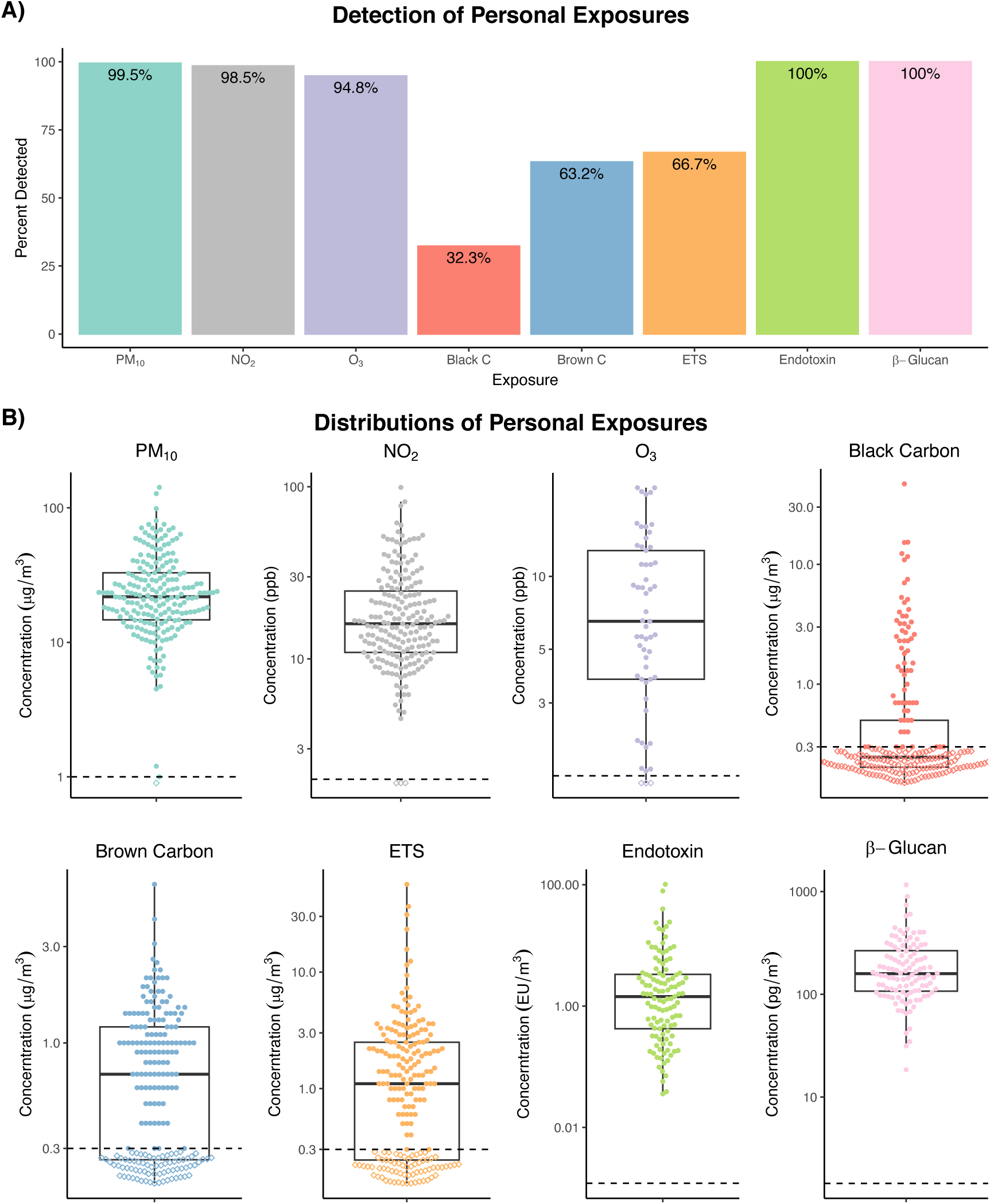
A) Detection of exposures using personal monitoring. B) Box and whisker plots of personal exposures. Boxes indicate the first, second, and third quartiles of the distribution. Whiskers extend to 1.5 times the inter-quartile range. Dashed lines indicate lower limits of detection.

**Table 2:** Description of personal exposures by season. N(%) or median [IQR].

| Exposure | LOD |  | Overall<br>(N =219) | Fall<br>(N =60) | Spring<br>(N =42) | Summer<br>(N =63) | Winter<br>(N =54) |
| --- | --- | --- | --- | --- | --- | --- | --- |
| **O3** | 1.5 ppb | Below LOD<br>Above LOD<br>Median<br>[IQR] | 3 (5.2)<br>55 (94.8)<br>6.4<br>[3.8, 12.5] |  |  |  |  |
| **NO2** | 2 ppb | Below LOD<br>Above LOD<br>Median<br>[IQR] | 3 (1.5)<br>196 (98.5)<br>16.0<br>[10.9, 24.8] | 1 (2.0)<br>49 (98.0)<br>15.2<br>[10.7, 21.6] | 1 (2.7)<br>36 (97.3)<br>16.1 [10.9,<br>28.2] | 1 (1.7)<br>59 (98.3)<br>15.2<br>[9.8, 24.5] | 0 (0.0)<br>52 (100.0)<br>18.6<br>[13.0, 27.4] |
| **PM10** | 1 ug/m3 | Below LOD<br>Above LOD<br>Median<br>[IQR] | 1 (0.5)<br>208 (99.5)<br>21.8<br>[14.7, 32.9] | 0 (0.0)<br>58 (100.0)<br>23.3<br>[16.1, 31.6] | 0 (0.0)<br>38 (100.0)<br>21.3 [15.8,<br>39.6] | 0 (0.0)<br>61 (100.0)<br>21.6 [14.6,<br>35.9] | 1 (1.9)<br>51 (98.1)<br>22.6<br>[13.5, 31.5] |
| **Black Carbon** | 0.3 mg/m3 | Below LOD<br>Above LOD<br>Median<br>[IQR] | 136 (67.7)<br>65 (32.3)<br>0.3<br>[0.3, 0.5] | 42 (73.7)<br>15 (26.3)<br>0.3<br>[0.3, 0.3] | 30 (88.2)<br>4 (11.8)<br>0.3 [0.3,<br>0.3] | 28 (47.5)<br>31 (52.5)<br>0.3 [0.3,<br>2.2] | 36 (70.6)<br>15 (29.4)<br>0.3 [0.3,<br>0.4] |
| **Brown Carbon** | 0.3 mg/m3 | Below LOD<br>Above LOD<br>Median<br>[IQR] | 74 (36.8)<br>127 (63.2)<br>0.7<br>[0.3, 1.2] | 12 (21.1)<br>45 (78.9)<br>0.9<br>[0.6, 1.2] | 6 (17.6)<br>28 (82.4)<br>0.8 [0.4,<br>1.4] | 38 (64.4)<br>21 (35.6)<br>0.3 [0.3,<br>0.9] | 18 (35.3)<br>33 (64.7)<br>0.6<br>[0.3, 1.1] |
| **ETS** | 0.3 mg/m3 | Below LOD<br>Above LOD<br>Median<br>[IQR] | 67 (33.3)<br>134 (66.7)<br>1.1<br>[0.3, 2.5] | 7 (12.3)<br>50 (87.7)<br>1.4<br>[0.8, 2.6] | 8 (23.5)<br>26 (76.5)<br>2.3 [0.4,<br>3.5] | 36 (61.0)<br>23 (39.0)<br>0.3 [0.3,<br>1.3] | 16 (31.4)<br>35 (68.6)<br>1.1<br>[0.3, 2.5] |
| **Endotoxin** | 0.0012 EU/m3 | Median<br>[IQR] | 1.4<br>[0.4, 3.3] | 2.1<br>[0.7, 3.2] | 0.8<br>[0.2, 2.6] | 1.6<br>[0.5, 3.8] | 1.6<br>[0.5, 3.4] |
| **Glucan** | 1.447 pg/m3 | Median<br>[IQR] | 158.7<br>[107.5,<br>265.5] | 178.2<br>[119.3,<br>238.0] | 175.6<br>[109.3,<br>302.8] | 159.9<br>[107.0,<br>241.1] | 144.8<br>[97.3,<br>201.5] |

Regarding PM_10_ constituents, all samples had detectable endotoxin and 𝛽-glucan, and wide variation in levels of these exposures, with endotoxin ranging from 0.04 to 101.3 EU/m^3^ and 𝛽-glucan levels ranging from 18.5 to 1,162 pg/m^3^. Detection rates were lower for other PM_10_ constituents with only 32.3%, 63.2%, and 66.7% of samples having detectable BC, BrC, and ETS respectively. BC, BrC and ETS ranged from <0.3 to 46.9, <0.3 to 6.1, and <0.3 to 56.6 𝜇g/m^3^, respectively.

### Seasonal variation in pollutant exposures

To understand if seasonal fluctuations could explain the wide variation in exposures levels, we compared exposure levels for each pollutant between seasons (STable 1). Personal PM_10_ levels did not significantly vary by season (Figure 3). In contrast, NO_2_ levels were significantly lower in the summer compared to winter (20.1% lower, CI: 32.3% to 5.8% lower; FDR = 0.019); however, there was still substantial overlap in the distribution of NO_2_ levels between seasons.

**Figure 3.**
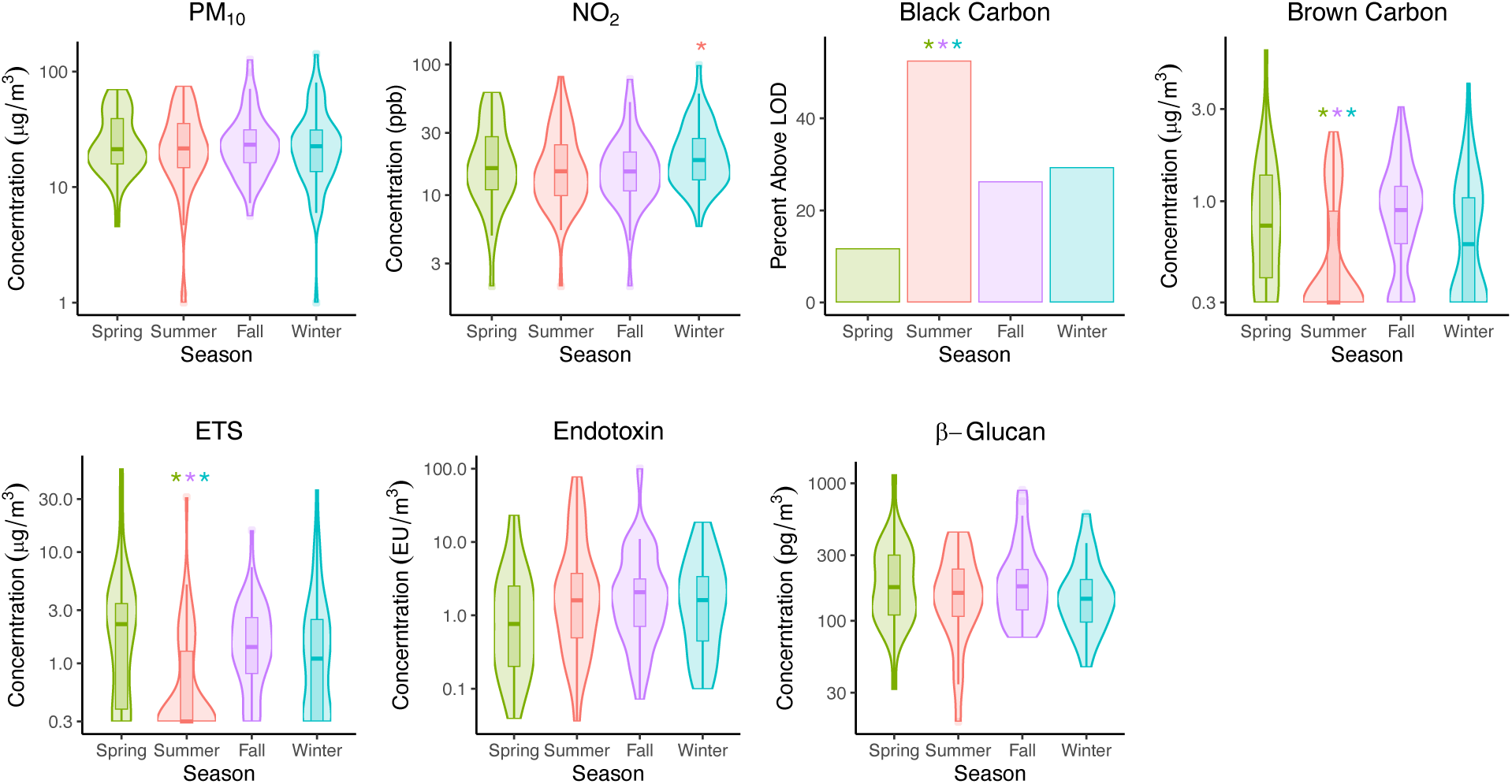
Distributions of personal exposures by season. Boxes indicate the first, second, and third quartiles of the distribution. Whiskers extend to 1.5 times the inter-quartile range. Stars indicate significant differences between seasons (FDR adjusted p-value < 0.05).

While total PM_10_ did not significantly differ between seasons, there were seasonal differences in the levels of several PM_10_ constituents. ETS and BrC levels were significantly lower in the summer compared to all other seasons. The largest seasonal difference was between summer and fall, with ETS and BrC exposure levels being 77.3% (CI: 87.2% to 59.6% lower; FDR<0.0001) and 61.5% (CI: 73.9% to 43.1% lower; FDR<0.0001) lower in the summer, respectively. While at relatively low levels, BC followed a different pattern, with significantly higher odds of detection in the summer months compared to all other seasons (Odds Ratios: Summer v Spring 7.48 (2.45 to 22.82; FDR = 0.0009); Summer v Winter 2.71 (1.26 to 5.82; FDR = 0.019); Summer v Fall 2.99 (1.36 to 6.56; FDR = 0.015)). Endotoxin and 𝛽-glucan levels did not significantly differ between seasons.

### Person-level exposures vary between children and within children over time

Since there was substantial overlap in the distribution of exposures between seasons, we next considered whether the large range in observed values was more reflective of differences in exposure environments encountered by different participants (between participant variation) or changes in the environment encountered by the same participants (within participant variation). Examining this, we found large differences in average PM_10_ levels between participants, with some participants having average exposures less than 10 𝜇g/m^3^ and others having average exposures nearly 10-fold higher (Figure 4A).

**Figure 4.**
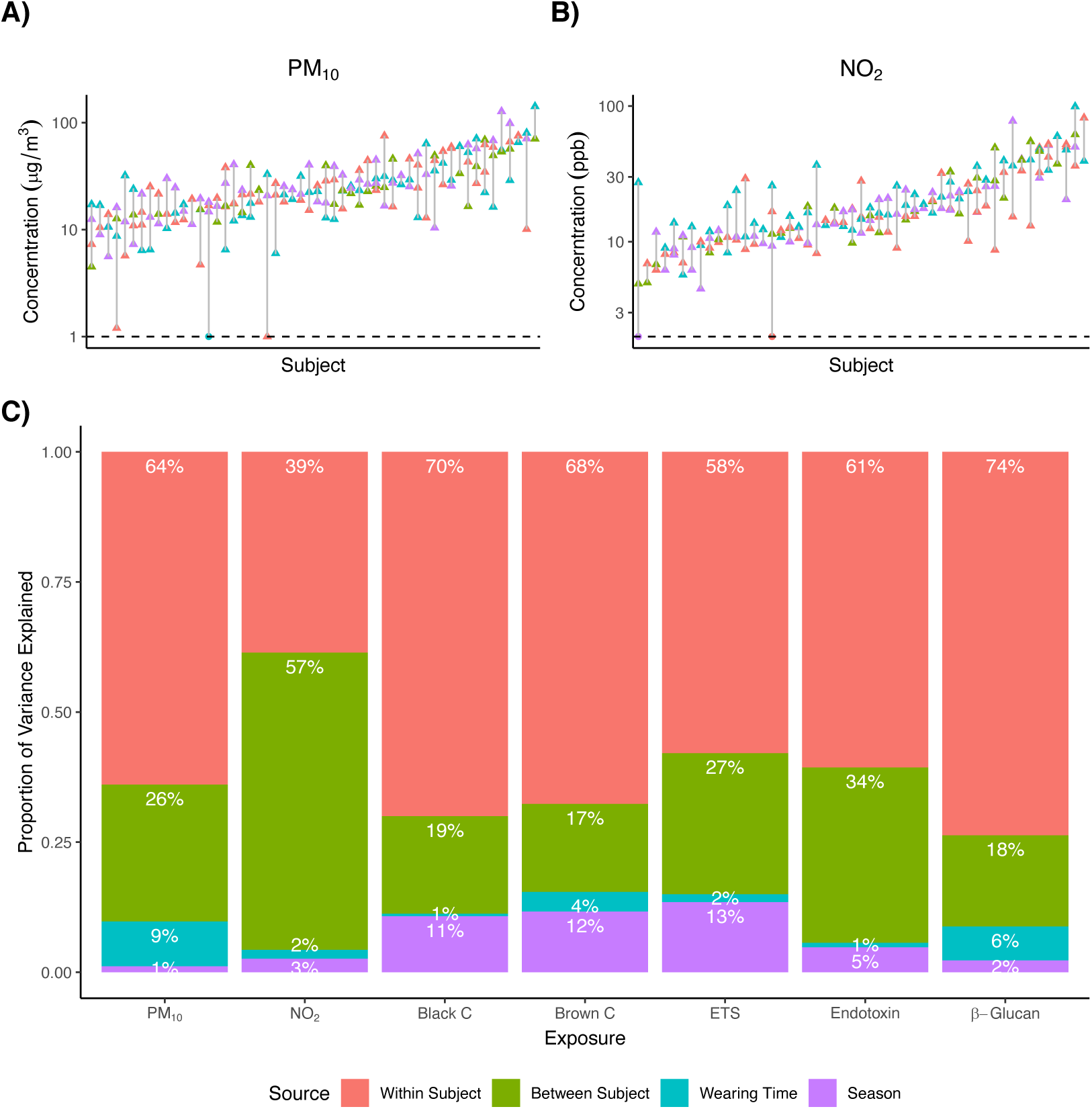
A) Dot plot of personal PM_10_ measures for subjects with at least 2 PM_10_ observations. Each column in the plot represents a subject. Dots indicate individual PM_10_ measures and lines indicate the range of personal PM_10_ for a subject. B) Dot plot of personal NO_2_ measures for subjects with at least 2 NO_2_ observations. C) Sources of variability in personal exposure data. Variation that can be explained by season, percent of time the participant wore the monitor, between subject differences, and the remaining within subject variability are shown in purple, blue, green and pink, respectively.

However, there was also wide variation in PM_10_ levels within individual participants, with an average 2.8-fold difference between a participant’s highest and lowest PM_10_ measurement. Similarly, for NO_2_, the lowest average exposure for a participant was 5.9 ppb compared to 57.7 ppb for the participant with the highest average exposure (Figure 4B). For NO_2_ there was an average 2.0-fold difference between a participant’s highest and lowest measurement. Similar patterns can be seen for other exposures (SFigure 1).

To quantify this more formally, we used linear mixed effects models to estimate the proportion of variability in personal exposures that could be attributed to seasonal variation, differences in wearing compliance (measured as the percent of waking hours during which the participant wore the monitor), between participant differences, and the remaining within participant variability (Figure 4C). Season explained a relatively small proportion of the variability in exposure levels, ranging from 1% for PM_10_ to 14% for ETS, as did wearing time (range: 1% for BC and endotoxin to 9% for PM_10_). Between participant differences accounted for a more substantial proportion of the variability in personal exposures, ranging from 17% for BC to 59% for NO_2_, suggesting that even though participants live in the same general geographic location, there are still large differences in the exposure levels experienced by these participants. To understand the drivers of between participant variability in exposure levels, we tested for association between participant and household characteristics and personal exposures. Neither age, sex, race-ethnicity, nor BMI were significantly associated with any of the exposures; however, there were associations between socio-economic factors and personal PM_10_ (STable 1). Children with parents with a high school education or less had 1.46-times higher exposure to PM_10_ (CI:1.12-1.92, FDR=0.046), and children from households with annual incomes less than $20,000 also had 1.45-times higher exposure to PM_10_ (95% CI:1.13-1.88, FDR=0.036).

Finally, we assessed the influence of within-participant variability on variation in exposure levels. For most exposures we found within-participant variability was a substantial contributor to variation in exposure levels, ranging from 38% for NO_2_ to 73% for BC (Figure 4C). This within participant variability could reflect longitudinal variation in microenvironments encountered by individual participants, changes in activity levels or other behavioral changes, as well as technical variation.

### Agreement between personal, community air quality, and in-home exposure measurements is limited

#### Personal vs In-Home Monitoring

A subset of 49 pre-pandemic participants had in-home monitoring for NO_2_, PM_10_ and PM speciation from stationary monitors placed in the participant’s bedroom during the same time period as the personal monitoring, allowing us to assess correlation between personal and in-home monitoring detection levels. In-home measurements of PM_10_ and NO_2_ were strongly associated with personal exposures, particularly for NO_2_ (Pearson correlation: r = 0.7, r=0.96 for PM_10_ and NO_2_ respectively, Figure 5A). However, despite the high level of correlation between in-home and personal measurements, PM_10_ was an average of 47.6% higher when measured with the personal monitor (limits of agreement: 45.6% lower to 300.7% higher, STable 2). For NO_2_, exposure levels were an average of 8.1% higher when measured with the personal monitor compared to the in-home monitor (95% limits of agreement: 33.4% lower to 75.3% higher).

**Figure 5.**
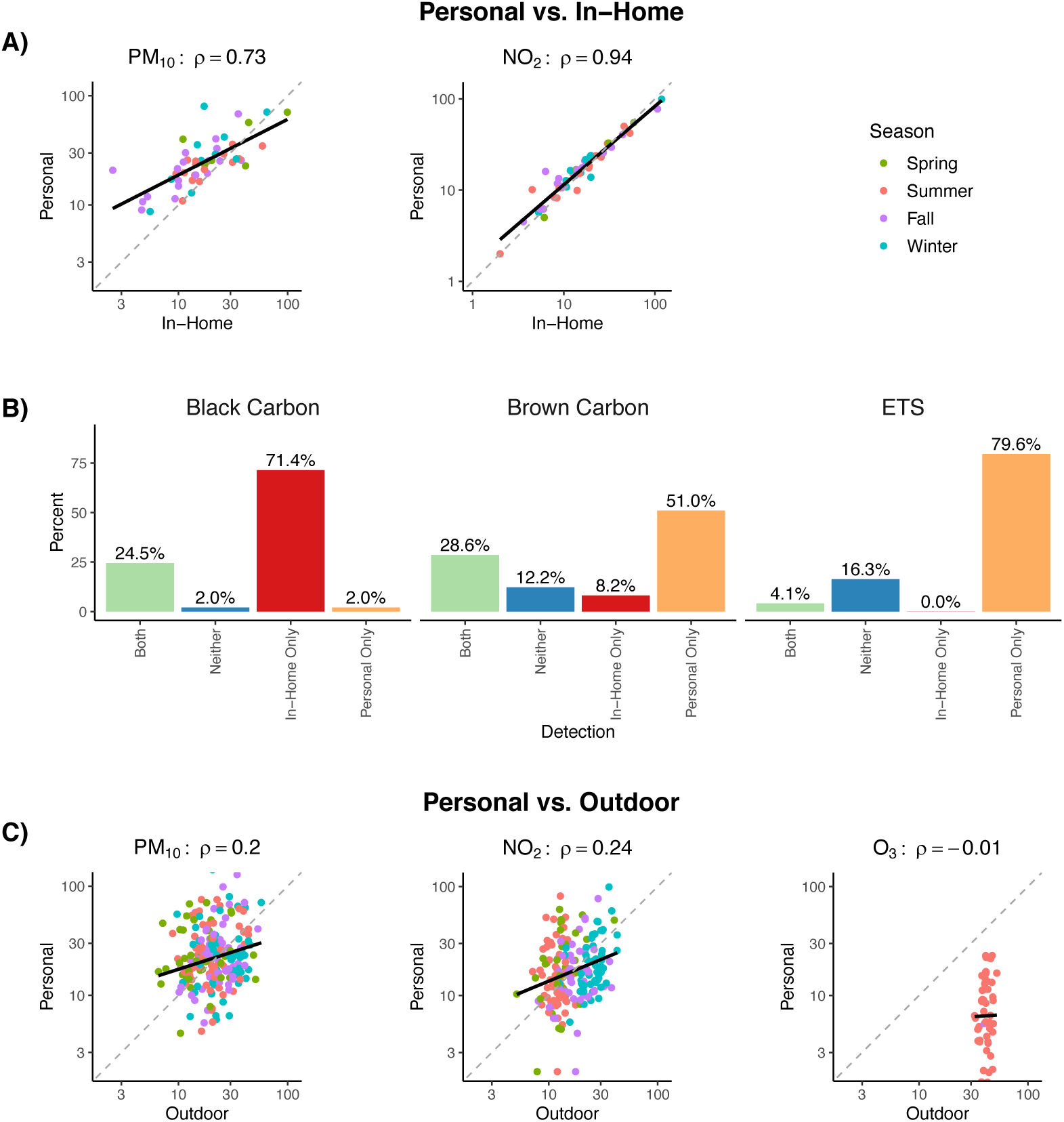
A) Scatterplots and Spearman correlations comparing personal to community outdoor exposure measurements, colored by season. B) Scatterplots and Spearman correlations comparing personal to in-home exposure measurements, colored by season. C) Detection of PM_10_ constituents by personal and in-home monitors. Green bars indicate the exposure was detected by both monitors, blue bars indicate the exposure was detected by neither monitor, red bars indicate the exposure was detected by the in-home monitor only, and orange bars indicate the exposure was detected with the personal monitor only.

Examining PM constituents by detection (yes/no, Figure 5B), BrC and ETS were more likely to be detected with the personal monitor, despite the fact that the in-home monitor had a lower LOD (0.1 ug/m^3^) than the personal monitor (0.3 ug/m^3^). BrC was detected in 79.6% of personal monitoring sessions, compared to 36.8% in-home sessions, and ETS was detected in 83.7% of personal monitoring sessions vs. 4.1% of in-home sessions, suggesting that smoke exposure may be occurring outside of the home or in other rooms of the home, outside of the child’s bedroom. BC was more often detected using the in-home monitor (94.9% vs 26.5%), though this may be driven in part by the lower LOD of the in-home monitor. Together these data suggest that while there is some level of correlation between in-home pollutant levels and personal exposure levels, levels of exposures and even exposure occurrence in general is greatly underestimated using home monitoring.

#### Personal vs. Community Monitoring

We similarly examined the correlation between community monitoring and person-monitor levels of PM_10_ and NO_2_. These analyses found personal exposure levels of PM_10_ and NO_2_ were only weakly correlated with the detected outdoor levels (r = 0.20, 0.24 for PM_10_ and NO_2_, respectively, Figure 5C). On average, personal exposures to PM_10_ were 10% higher than community outdoor levels (95% limits of agreement: -68.5% lower to 285.1% higher), and personal NO_2_ was 0.5% higher than outdoor measurements (95% limits of agreement: -76.6% lower, 323.5% higher). There was no association between personal and community outdoor O_3_ measurements (r=-0.01). While measured outdoor O_3_ levels were all within a relatively narrow range (25.3 to 51.7 ppb), personal exposures had greater variation, ranging from 1.5 to 23.3 ppb. On average, personal O_3_ measurements were 84.7% lower than community outdoor measurements (95% limits of agreement: 96.8% lower to 26.2% lower). Together, these data suggest true personal exposures to PM_10_, NO_2_, and O_3_ are poorly estimated by community monitor measured levels.

### Person-level exposures are associated with asthma exacerbation

Examining the relationship between personal exposures and asthma severity, we found exposure levels did not significantly differ between participants with and without well controlled asthma, based on cACT and ACT scores (STable 3). We also evaluated the associations of exposures with lung function measures (STable 4). We did not observe any statistically significant associations between exposures and pre-bronchodilator spirometry or change post-bronchodilator. Community air quality and indoor stationary monitor exposure levels were not significantly associated lung function or asthma control, although indoor measurements were only available on 49 subjects (SFigures 2-4).

We next investigated the relationship between personal exposure levels and asthma exacerbation (Figure 6, STable 3). We found participants that reported an unscheduled healthcare visit for asthma in the 60 days prior to their exposure assessment had 1.35, 1.57 and 2.15-times higher levels of PM_10_ (CI: 1.08 to 1.69; FDR =0.037), BrC (CI: 1.09 to 2.26; FDR =0.040) and ETS (CI: 1.36 to 3.42; FDR =0.009), respectively (STable 3). Similarly, those requiring a course of systemic corticosteroids for asthma in the 60 days prior to their assessment had 1.98 and 2.21-times higher levels of BrC (CI: 1.43 to 2.37; FDR =0.0003) and ETS (CI: 1.25 to 3.91; FDR =0.026), as well as 2.04-times higher endotoxin (CI: 1.14 to 3.68; FDR =0.045). In addition, participants that were hospitalized for asthma in the 18 months prior to enrollment had 2.6-times higher summer O_3_ exposure (CI: 1.69-4.00; FDR = 0.0001). Community air quality and indoor stationary monitor exposure levels did not significantly differ between subjects with and without exacerbation (SFigure 2). Together, these results show personal pollutant exposures are a strong predictor of asthma exacerbation outcomes.

**Figure 6.**
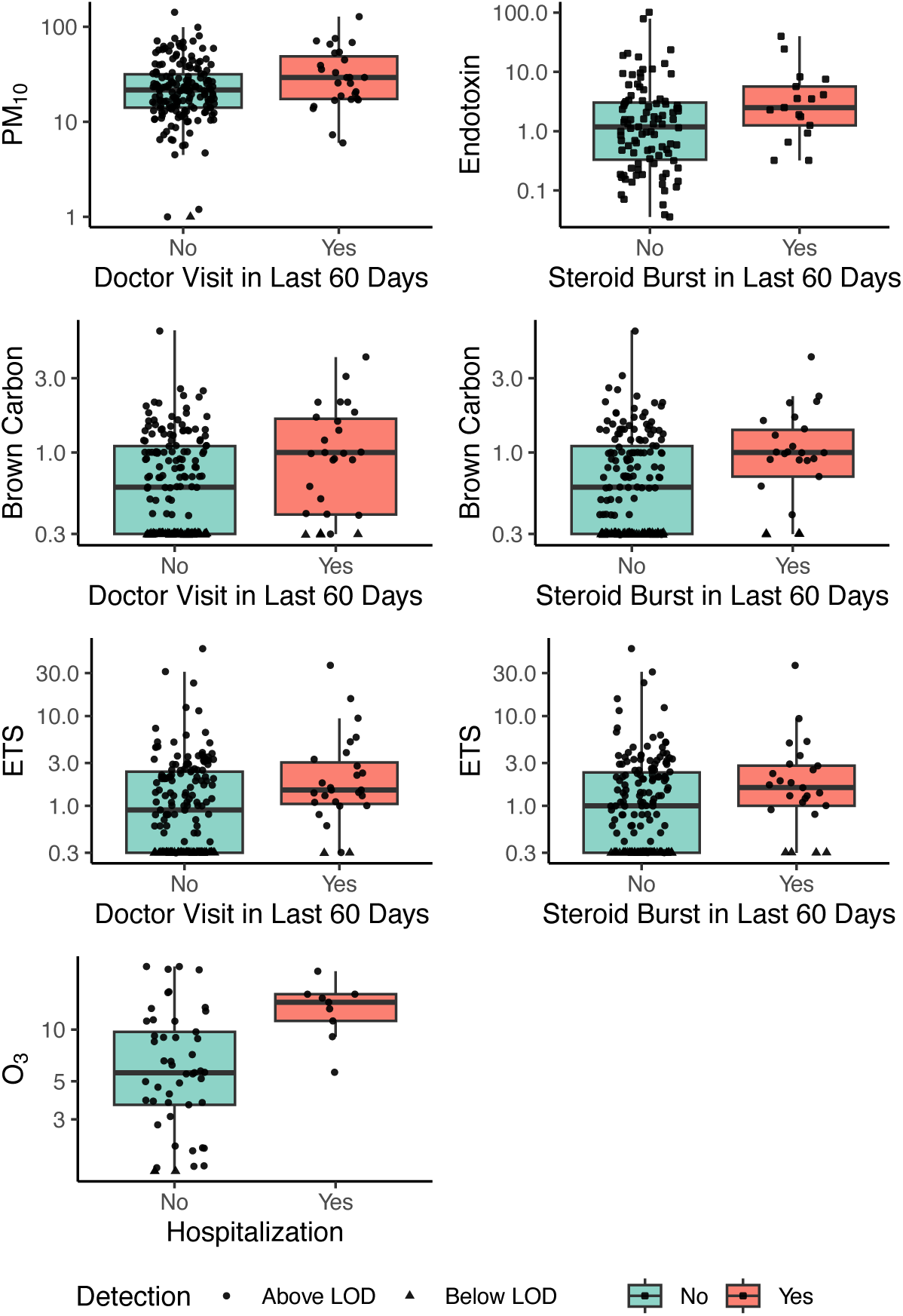
Box and whisker plots of personal exposures by asthma exacerbation characteristics.

## DISCUSSION

Although multiple epidemiologic studies have associated regional spikes in airborne pollutant levels with increases in acute respiratory events, including asthma exacerbations, the precise relationships and degree of effects is likely obscured by variation in personal exposure levels. Here, we leveraged cutting-edge, wearable monitors, capable of measuring breathing zone pollutant exposure levels, to investigate the uniqueness of these measurements and their added value in exploring exposure-outcome relationships. Our findings suggest that community monitoring of PM_10_, NO_2_, and O_3_ is a poor proxy for exposures experienced by children with EP-asthma. Moreover, we find that although in-home monitoring is correlated with personal exposure to PM_10_ and NO_2_, in-home detection of some of the most important, asthma-provoking PM constituents is unrelated to the actual breathing zone exposures children experience. Excitingly, we leveraged these unique breathing zone exposure measurements, to show PM_10_ levels in general, as well as BrC, ETS, and endotoxin levels are all associated with childhood asthma exacerbations.

A key assumption underlying the use of community air quality monitoring data in studies investigating the impact of air pollution on asthma outcomes is that all participants in a geographical region have similar exposures that are accurately characterized by outdoor pollution levels. However, similar to other studies,^63^ in the ENIGMA cohort community outdoor PM_10_, NO_2_ and O_3_ levels did not accurately reflect an individual child’s exposures. In addition, though participants were from a relatively small geographical region, we observed substantial variation in personal exposures, with an approximately 10-fold difference in PM_10_ and NO_2_, and 16-fold difference in O_3_ levels, between participants with the highest and lowest exposures. While PM_10_ was associated with socioeconomic factors, differences in exposures between participants are also likely driven by behavioral factors, such as the amount of time spent outdoors, their proximity to major roads and other strong outdoor air pollution sources, and differences in indoor microenvironment concentrations at home and in school^64^.

While indoor monitors placed in participants’ bedrooms were associated with personal measures, there were important differences. On average, personal PM_10_ measurements were approximately 50% higher than in-home measurements, suggesting that indoor stationary monitors substantially underestimate PM_10_ exposure. In particular, ETS and BrC were more likely to be detected with personal monitors, suggesting tobacco smoke exposure occurred in other areas of the home (outside the child’s bedroom) or outside of the home, limiting the utility of in-home monitors in assessing these exposures. Interestingly, BC was more likely to be detected with in-home monitors. The main source of indoor BC is infiltration of outdoor pollution from diesel combustion, which is more pronounced in homes without air conditioning that rely on open windows for cooling and ventilation during the summer months.^65^ In addition, many of Denver’s residential neighborhoods flank interstates and highways, which could cause elevated levels of BC in some households.^66^

While personal monitoring provided a 72-hour snapshot of the exposures experienced by individual participants, personal PM_10_ and several PM constituents were associated with exacerbation outcomes over a broader timeframe, suggesting that short-term monitoring may provide information about typical exposure levels experienced by a participant over a longer period. Despite this, we observed within participant variability in exposure levels beyond what could be explained by seasonal fluctuations. This variation in exposure measurements within a participant suggests that repeated or longitudinal personal monitoring may be valuable to accurately capture a participant’s exposure levels.

Similar to other more recent studies using personal measurements, we did not find associations between personal exposures and short-term changes in lung function or daily asthma control (ACT).^67–69^ While Delfino *et al*^70^ observed FEV_1_ was associated with PM_2.5_ and NO_2_ in children with asthma, these findings were only significant among those who did not use bronchodilators. The broad use of asthma control medications among children with EP asthma may explain, in part, why lung function was not associated with personal exposures in the ENIGMA cohort. However, personal PM_10_ and PM_10_ constituents, including ETS, BrC and endotoxin, were associated with exacerbation outcomes, including systemic corticosteroid courses and unscheduled healthcare visits for asthma. While previous studies have found associations between PM_10_ and PM_2.5_ and asthma exacerbations,^71–78^ our study provides insights into more specific PM constituents impacting asthma. While numerous studies have confirmed the deleterious effect of ETS exposure on asthma outcomes,^79–83^ the role of BrC is less fully understood. BrC is emitted from burning biomass, including forests fires, residential heating with coal and wood and cooking, from “biogenic release of fungi, plant debris, and humic matter,” as well as from secondary atmospheric reactions.^84, 85^ Few studies have investigated the specific role of BrC in respiratory outcomes, although outdoor BrC levels have been associated with increased risk of acute respiratory infection in children.^86^ In addition, wildfires, a source of BrC, have been associated with increased rates of emergency department visits and hospitalization for asthma, with some evidence suggesting that children are particularly vulnerable to wildfire associated exacerbations.^87–93^ Endotoxins, lipopolysaccharides and lipo-oligosaccharides from the outer cell wall of Gram-negative bacteria, have been shown to induce innate immune responses and inflammation in mouse studies,^94^ as well as in controlled human experiments.^95^ While early life endotoxin exposure has been associated with reduced risk of developing allergies and allergic asthma,^96^ personal and classroom endotoxin levels have been associated with increases in asthma symptoms and lung function among school age children with asthma^97, 98^ and endotoxin exposure was associated with doctor and emergency room visits for wheeze in the NHANES study.^99^

Despite these important findings, some study limitations should be noted. Namely, although our investigation of personal exposures in children with EP asthma, specifically from the Denver metro area, allowed us to assess the level of variability in exposures among asthmatic children in a single geographic region, it may limit the generalizability of our findings to other geographic areas or to children with less severe disease. In addition, since our cohort was of moderate size and longitudinal follow up was not available on all participants due COVID-19 pandemic related study disruptions, power to detect associations between exposures and asthma outcomes was reduced, particularly for exposures with high proportions of measurements below the LOD. Additionally, although our assessment of PM was limited to PM_10_ in order to collect sufficient sample for constituent analyses, we acknowledge a likely role for PM_2.5_ (particles less than 2.5 microns in diameter) exposures in driving asthma outcomes, as has been reported^70, 100–102^.

Despite these limitations, our study highlights how personal exposure monitoring can be used to accurately characterize breathing zone pollutant exposure levels, providing a more nuanced understanding of the PM constituents contributing to asthma exacerbation.

## Supporting information

STable

## Data Availability

All data produced in the present study are available upon reasonable request to the authors

## ACKNOWLEDGEMENTS

ENIGMA was funded by the National Heart, Lung and Blood Institute, National Institutes of Health, grant number 5P01HL132821.

## ABBREVIATIONS

ACT: Asthma Control Test
BC: black carbon
BrC: brown carbon
cACT: Childhood ACT
CASI: Composite Asthma Severity Index
EP: exacerbation-prone
ETS: environmental tobacco smoke
FDR: false discovery rate
IQR: interquartile range
LOD: limit of detection
NO_2_: nitrogen dioxide
O_3_: ozone
PM_10_: particulate matter less than 10 microns in diameter
PM_2.5_: particulate matter less than 2.5 microns in diameter
ppb: parts per billion

## Notes

### Competing Interest Statement

Camille M. Moore, Elizabeth A. Secor, Allison M. Schlitz, Kristy L. Freeman, Jamie L. Everman and Tasha E. Fingerlin have no financial disclosures or conflicts of interest to report. Jonathan Thornburg serves as the treasurer for the International Society of Exposure Science. Andrew H. Liu has received grants paid to Children's Hospital Colorado from ResMed and OM Pharma, consulting fees paid to the University of Colorado from ThermoFisher Scientific, and research equipment from ResMed/Propeller Health and Revenio. Andrew H. Liu has also served on data safety monitoring boards for AstraZeneca. Max A. Seibold received an honorarium for lecturing at the Colorado Allergy and Asthma Society.

### Author Declarations

The Colorado Multiple Institutional Review Board gave ethical approval for this work.

