## Supplement for "Breathing zone pollutant levels are associated with asthma exacerbations in high-risk children"

**Supplementary Methods**

***Personal Exposure Monitoring***

Personal exposure monitoring was performed for approximately 72 hours prior to all scheduled study visits performed through May 2022, with a total of 81 participants completing 1 or more personal monitoring sessions. We used two approaches to collect environmental exposure samples. Pre-COVID, field personnel visited the home to deploy the RTI MicroPEM™ (Research Triangle Park, NC) and Ogawa passive dosimeters (Ocala, FL). All devices were placed in a belt bag that was placed diagonally across the chest such that the monitor inlet was within the breathing zone i.e., within 10 inches of the nose and mouth.[1, 2] We instructed the participant to wear the pouch as much as possible during the 72-hour monitoring period. Acceptable times to keep the pack nearby but not worn were while sleeping, showering or bathing, sports or similar activities where the pack would interfere with their movement. Field personnel retrieved the samples after the monitoring period and stored them at 4°C until shipped to RTI for analysis. In May 2020, we switched to a direct shipping deployment because of COVID. All devices and the belt bag were shipped from RTI directly to the participants home along with detailed photographic instructions how to place the samples in the belt bag and how to start the MicroPEM. The participant returned the samples to RTI after the monitoring period was completed.

The MicroPEM is a lightweight (~230 g), quiet (~ 45 dB) particulate matter sampling device designed to be worn by children and adults. The device sampled $PM_{10}$ at 0.4 Lpm for real-time nephelometer measurements every 10 seconds and subsequent collection on a 25 mm, 3 mm pore PTFE filter (Zefon International, Ocala, FL). Embedded temperature and relative humidity sensors automatically corrected the nephelometer concentration [3]. A 3-axis accelerometer measured children's activity each second to assess wearing compliance [4]. A rolling standard deviation of the accelerometer vector sum greater than 0.007 signified the child wore the MicroPEM for that 1-minute time interval. Wearing compliance was calculated number of minutes per day worn divided by the difference between the total monitoring time (minutes) minus their time not wearing the monitor but with it adjacent to them. Examples of this latter criterion are sleeping, at school, or showering.

A MicroPEM filter was valid if the sample duration was longer than 60 hours. Gravimetric analysis consisted of triplicate weights pre- and post-sample collection conducted on a Mettler UTM2 digital balance (Toledo, OH) inside a constant temperature (21°C) and relative humidity (35%) chamber [5]. The method detection limit for this study is 1.0 $\mu g/m^3$. A mean blank correction factor of 1.7 $\mu g$ developed from 28 laboratory blanks was applied to all gravimetric mass measurements. The 10 second nephelometer measures were gravimetrically corrected then averaged to a 1-minute measures. Black carbon (BC), brown carbon (BrC) and environmental tobacco smoke (ETS) mass on the filter were measured with a custom, 6-wavelength integrating sphere transmissometer [6]. The BC, BrC, and ETS method detection limits are 0.3 $\mu g/m^3$. Filters were then extracted for endotoxin and glucan analysis. The PTFE filters associated with the collection of nasal swabs were aseptically transferred to sterile 15 mL conical tubes using flame sterilized forceps. Two and a half milliliters of 1X PBS-T were added to each tube containing a filter. The tubes were shaken for 1 hour using a wrist action shaker (Burrell; Pittsburgh, PA). After shaking, 1 mL aliquot of each sample was transferred to an endotoxin/glucan free glass tube. A five-fold dilution (1:4) were prepared for each sample using the 1 mL aliquot. All were stored at 4°C and were used for the bioassays described below. The remaining extracts were stored in the conical tubes at -20°C. Both the endotoxin and glucan assays (Associates of Cape Cod; East Falmouth, MA) were set up according to RTI's Research Operating Procedures. Standard curves were generated for both assays using PBS-T buffer. Controls for the assays included blanks (PBS-T only) and a positive control containing either 0.5 EU/mL endotoxin standard or 25 pg/mL glucan standard. The endotoxin samples were analyzed undiluted and the glucan samples were analyzed at a five-fold dilution. The standards, controls, and samples were added to the endotoxin/glucan free 96 well plates in triplicate. The appropriate reaction buffer was added to each well and a kinetic assay was run measuring $OD_{405}$ at 37°C. Once completed, the standard curve was graphed on a log-log scale of the onset time versus the standard concentration. The correlation coefficient for the standard curve should have an absolute value of at least 0.980 to be valid. Unknown sample concentrations are determined using this graph. Samples outside of the standard curve range were designated as "OVER" or "BDL" (below the detection limit). Samples were considered valid if the CV for EU/mL or pg/mL for at least two wells was less than or equal to 15%. Detection limits were 0.0012 EU/$m^3$ and 1.45 units/$m^3$ for endotoxin and glucans, respectively.

$NO_2$ and $O_3$ Ogawa samples were collected on triethylamine and nitrite-coated filters, respectively. The filters were extracted with deionized water and the extracts analyzed by ion chromatography. The analytical detection limit was 24 ng for $NO_2$ and 0.015 □g for $O_3$. We applied analytical batch specific blank correction factors for $NO_2$ (Mean = 83.7 ng, Range: 18.9 – 341.3 ng, n = 86) and $O_3$ (Mean = 0.438 μg, Range: 0.136 – 0.822 μg, n = 25). The resulting mass was divided by the volume of air collected by diffusion calculated from equations provided by Ogawa[7] and the temperature and relative humidity measured by the MicroPEM.

***In-Home Environmental Exposure Assessment***

Pre-pandemic participants (n=49) also had in-home environmental monitors placed in their bedroom concurrent with their first 72-hour personal monitoring session. We collected $PM_{10}$ at 4L/min on PTFE filters (PEM™; MSP Corporation, Shoreview, MN) and monitored $NO_2$ (Ogawa), temperature, and humidity (HOBO®, Onset Computer Corporation, Bourne, MA). We analyzed samples for BrC, BC and ETS as described above.

***Outdoor Community Air Quality Data***

Hourly air quality data ($O_3$, $NO_2$, and $PM_{10}$) from the Denver Colorado Air Monitoring Program monitoring site were obtained from the Environmental Protection Agency's Air Quality System API for years 2018 through 2022. Sample durations of one hour were used to calculate the average exposure during the participant's start and end time for wearing their personal monitor.

#### *Clinical Assessments*

Clinical assessments included lung function measured by spirometry, asthma control, asthma severity, and questions on healthcare and medication usage in the previous 60 days. Spirometry was performed per American Thoracic Society/European Respiratory Society standards[8] with a clinical spirometer (KoKo® Legend, nSpire Health™, Longmont, CO, USA). Measurements were taken before and after four 90 μg puffs of albuterol to assess bronchodilator response. Global Lung Initiative reference equations were used for spirometry values.[9] Asthma control was assessed with the validated Asthma Control Test (ACT)[10] for participants ages 12 years and older (5-25 scale), and the Childhood ACT (cACT)[11, 12] for participants ages 6-11 years (0-27 scale). Scores from both tests have been combined in asthma research in children across the school age range (6 to 17 years), with scores >19 corresponding to well controlled asthma.[13] Asthma severity was scored with the validated Composite Asthma Severity Index (CASI) on a scale of 0 (best) to 20 (worst); this index combines day and night symptoms and albuterol use, daily controller therapy, lung function, and exacerbations, with scores < 4 indicating mild asthma.[14, 15]

#### *Statistical Methods*

*Person-Level Exposure Models.* We evaluated the association between personal exposures and participant and visit characteristics, including participant BMI, parental education level, household income, asthma control (cACT or ACT $\geq$ 20 vs. <20), hospitalization, systemic corticosteroid use, and health care visits, using censored regression models to account for observations below the limit of detection (LOD) with the ‘survival’ R package.[16, 17] We assumed a log-normal distribution for the exposure levels and used a robust ‘sandwich’ standard errors to account for repeated measures made on participants. As over 50% of BC measurements were below the LOD, we dichotomized BC levels into “Detected” vs. “Not Detected” and used generalized estimating equation (GEE) logistic regression models with the ‘geepack’ R package, instead of censored regression models.[18, 19] An exchangeable working correlation structure was used to account for repeated measures. Models controlled for age, sex, race-ethnicity, season and monitor wearing compliance, measured as the percent of awake time spent wearing the personal monitor. Models for $O_3$ were not adjusted for season as $O_3$ measurements were only collected in the summer. A Benjamini-Hochberg correction was applied to the p-values across the eight exposures to control the false discovery rate (FDR) at 5%.

*Intraclass Correlation*. To determine the proportion of variability in exposures that could be explained by between participant differences, we estimated intraclass correlation coefficients using linear mixed models adjusted for season with random intercepts to account for repeated measures on participants. Models were fit using a maximum likelihood approach to account for left censoring of observations below the LOD using the nlme procedure in SAS.

*Agreement Between Personal, Community Outdoor, and In-Home Monitors.* The ‘blandr’ R package[20] was used to calculate agreement between personal, outdoor, and in-home stationary monitoring measurements of $NO_2$ and $PM_{10}$ exposures. For participants with repeated measures, the first pairwise complete observation was taken in order to calculate agreement.

*Lung Function Models.* Pre-bronchodilator spirometry and the percent change in spirometry measures after bronchodilation were evaluated for association with the exposures using GEE models, assuming a normal distribution, using the ‘geepack’ R package. We used an exchangeable correlation structure to account for repeated measurements on participants.

Each exposure was evaluated in a separate model. The primary explanatory variable was the $\log_{10}$ of the exposure. The measured exposures had varying proportions of observations below the LOD. Depending on the amount of above LOD data available for an exposure, different analysis strategies were undertaken. For exposures where greater than 50% of observations were below the LOD (BC), the exposure was dichotomized into above and below the LOD. For exposures where less than 50% of observations were below the LOD ($PM_{10}$, $NO_2$, $O_3$, ETS, BrC), we imputed below LOD values as the $E(X \mid X < LOD)$ using methods described by Richardson and Ciampi.[21] Briefly, a Gaussian Tobit regression model,[22] which accounts for left-censoring of below LOD values, was fit to the log of the exposure values using the ‘survival’ R package.[16, 17] The estimated mean and standard deviation from this model was then used to calculate $E(X \mid X < LOD)$ using the R ‘truncnorm’ package. [23]

Models controlled for age, sex, race/ethnicity, wearing compliance, and season (with the exception of $O_3$ models). In addition, post-bronchodilation change models were adjusted for the corresponding baseline lung function measure.

### Supplementary Figures

**SFigure 1.** A) Dot plots of personal exposure measures for subjects with at least 2 observations. Each column in the plot represents a subject. Dots indicate individual exposure measures and lines indicate the range of personal exposure for a subject.

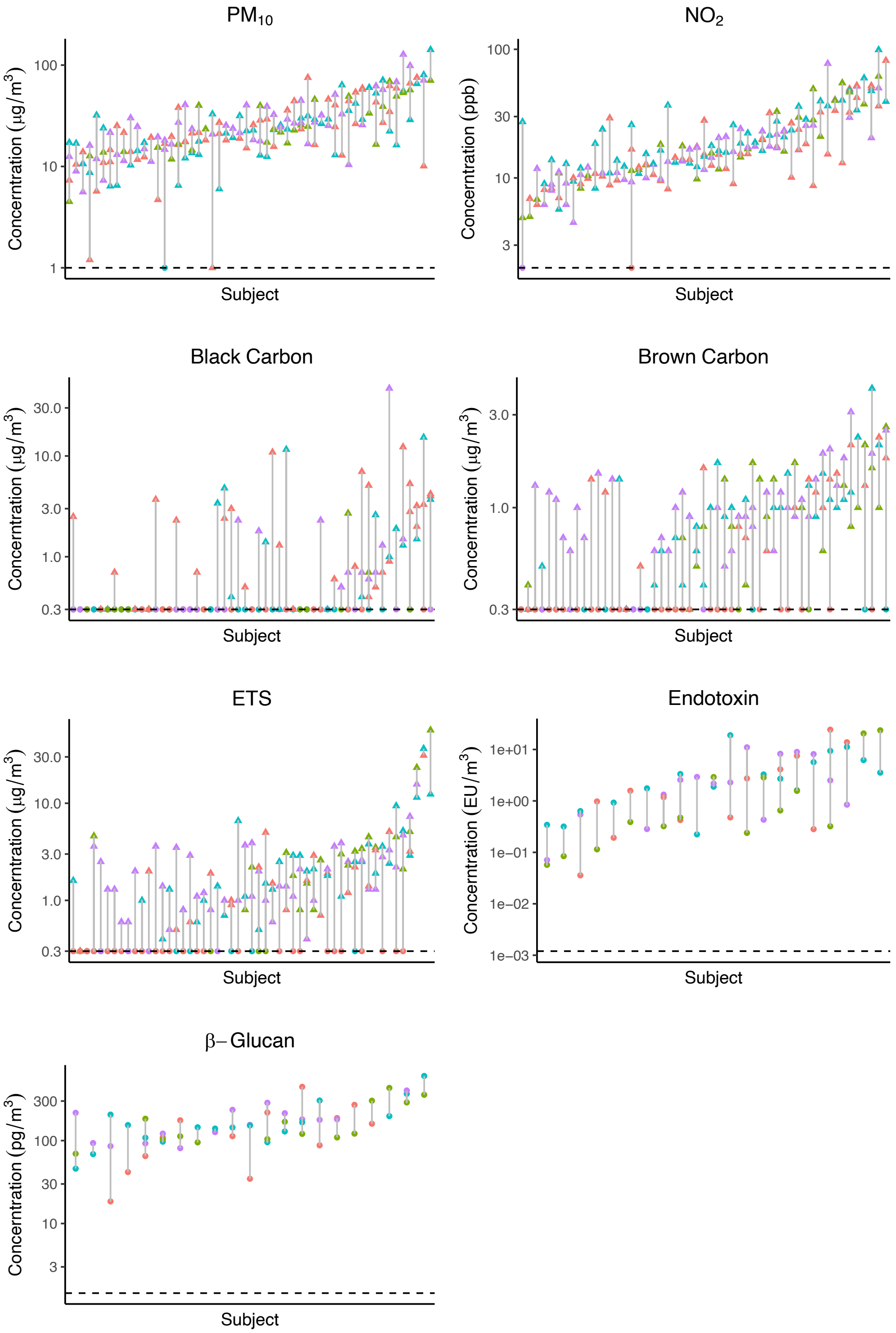

**SFigure 2.** Forest plots showing the fold difference in exposure levels between participants with and without an unscheduled asthma related healthcare visit in the prior 60 days, a course of systemic corticosteroids in the prior 60 days, hospitalization for asthma in the 18 months prior to enrollment, and ACT or cACT score less than 20. Stars indicate FDR-adjusted p-values < 0.05.

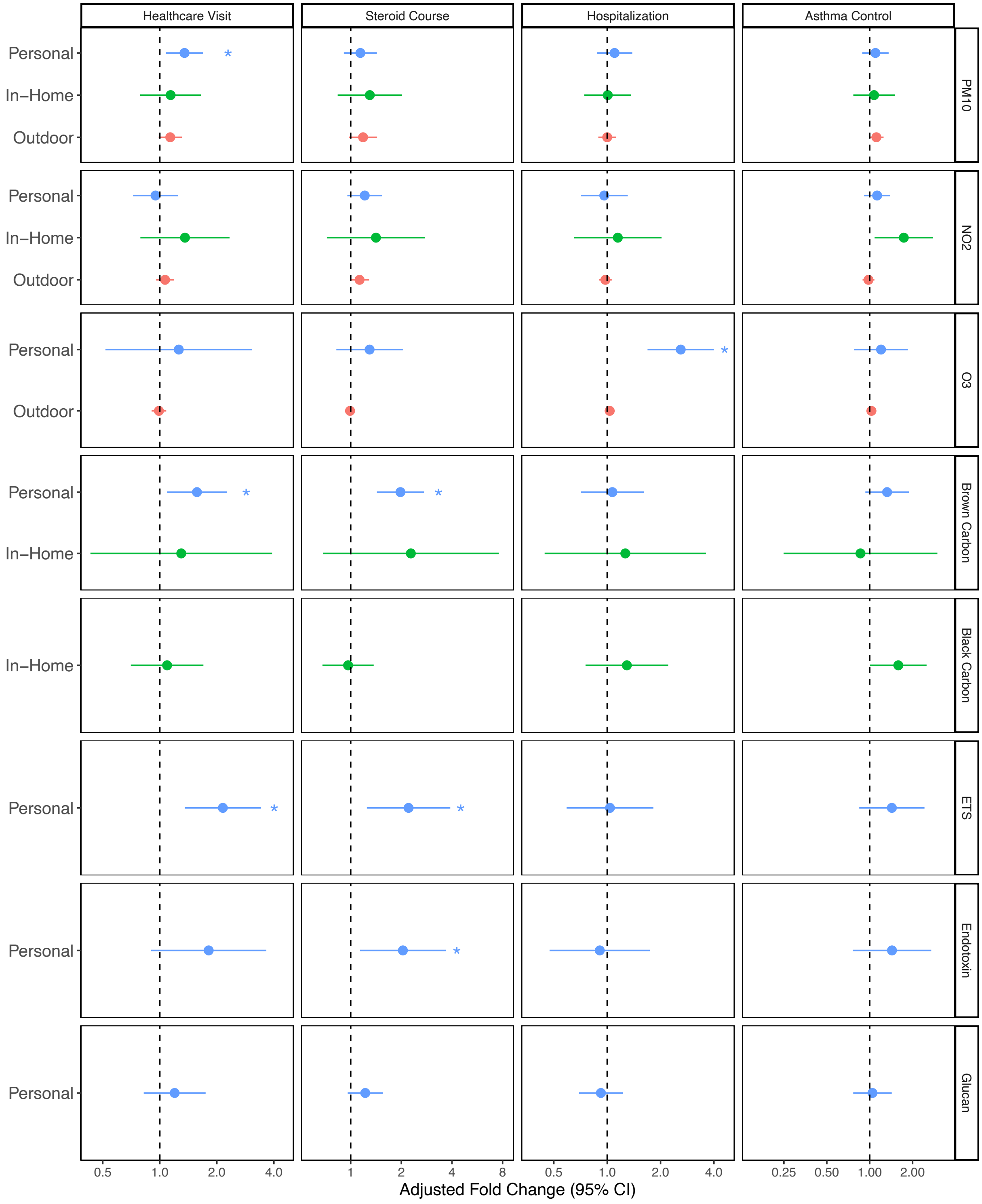

**SFigure 3.** Forest plots showing the estimated change in pre-bronchodilator spirometry (% predicted) for a 10-fold increase in exposure levels. Dots indicate the point estimate and the lines indicate the 95% confidence interval. Stars indicate FDR-adjusted p-values < 0.05.

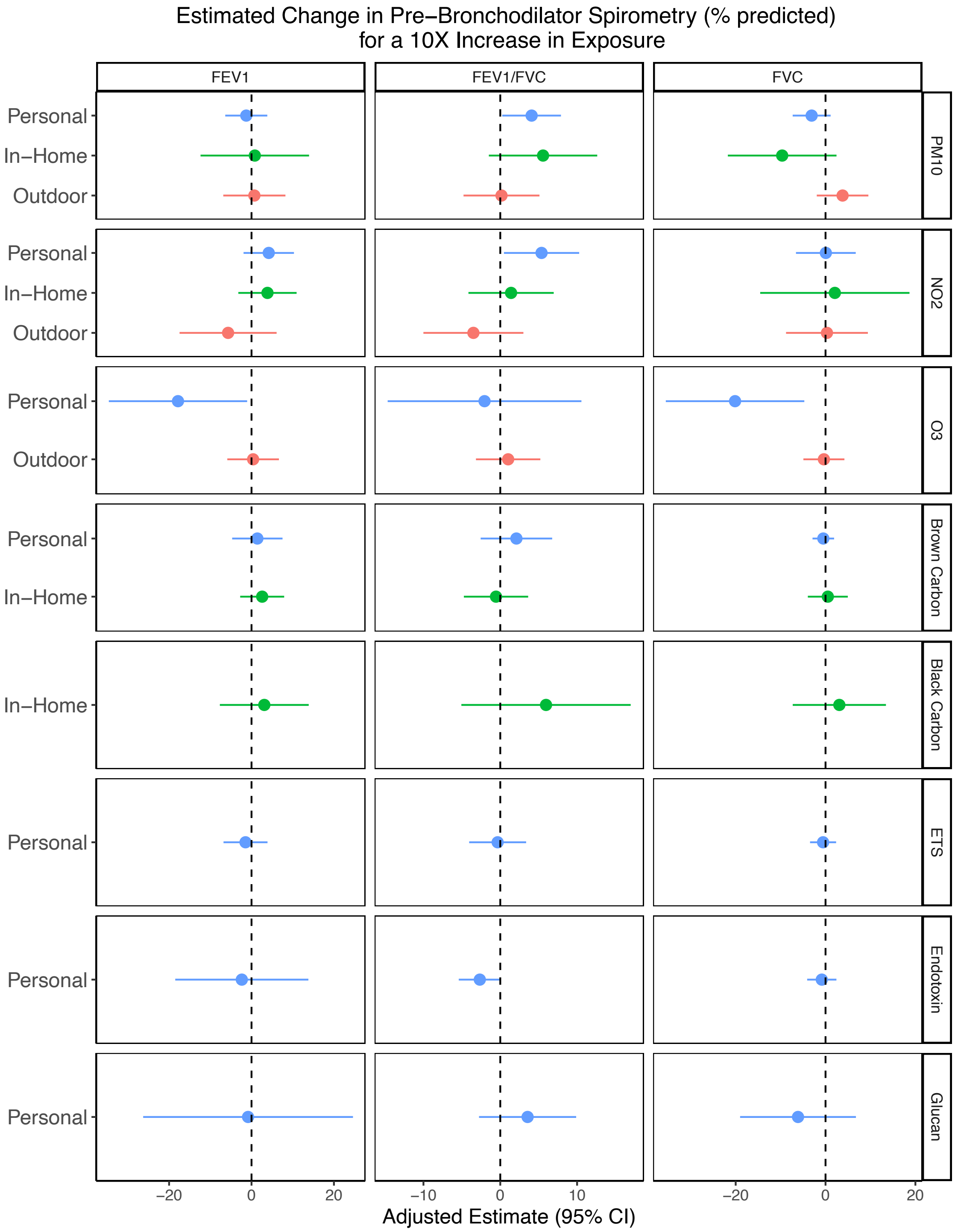

**SFigure 4.** Forest plots showing the estimated change in post-bronchodilator spirometry (% change from baseline) for a 10-fold increase in exposure levels. Dots indicate the point estimate and the lines indicate the 95% confidence interval. Stars indicate FDR-adjusted p-values < 0.05.

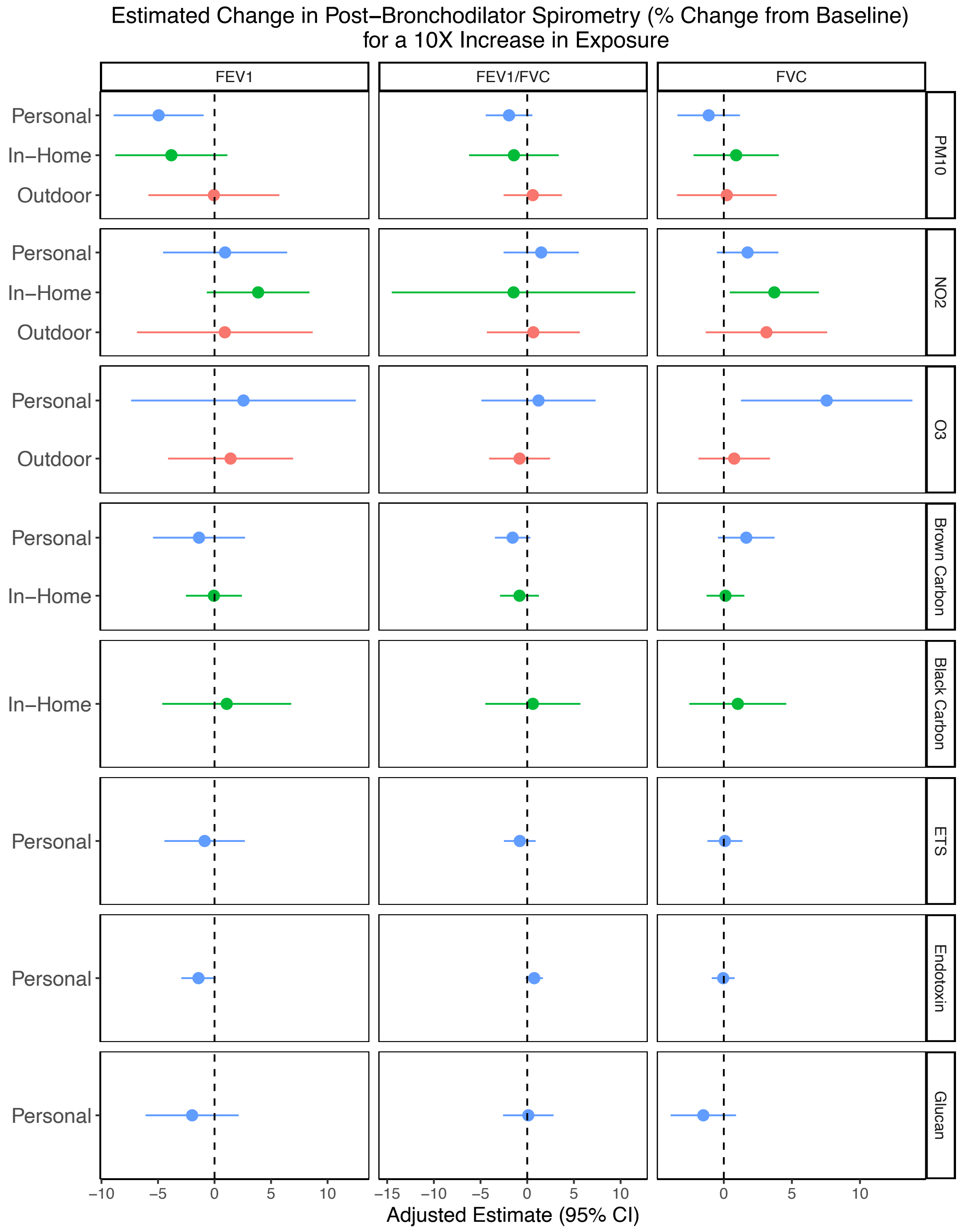

13. Pongracic JA, Krouse RZ, Babineau DC, Zoratti EM, Cohen RT, Wood RA, Khurana Hershey GK, Kercsmar CM, Gruchalla RS, Kattan M, Teach SJ, Johnson CC, Bacharier LB, Gern JE, Sigelman SM, Gergen PJ, Togias A, Visness CM, Busse WW, Liu AH. Distinguishing characteristics of difficult-to-control asthma in inner-city children and adolescents. J Allergy Clin Immunol. 2016;138(4):1030-41. doi: 10.1016/j.jaci.2016.06.059. PubMed PMID: 27720017; PMCID: PMC5379996.
14. Wildfire JJ, Gergen PJ, Sorkness CA, Mitchell HE, Calatroni A, Kattan M, Szefler SJ, Teach SJ, Bloomberg GR, Wood RA, Liu AH, Pongracic JA, Chmiel JF, Conroy K, Rivera-Sanchez Y, Busse WW, Morgan WJ. Development and validation of the Composite Asthma Severity Index-an outcome measure for use in children and adolescents. J Allergy Clin Immun. 2012;129(3):694-701. doi: 10.1016/j.jaci.2011.12.962. PubMed PMID: WOS:000301189300013.
15. Krouse RZ, Sorkness CA, Wildfire JJ, Calatroni A, Gruchalla R, Hershey GKK, Kattan M, Liu AH, Makhija M, Teach SJ, West JB, Wood RA, Zoratti EM, Gergen PJ. Minimally important differences and risk levels for the Composite Asthma Severity Index. J Allergy Clin Immun. 2017;139(3):1052-5. doi: 10.1016/j.jaci.2016.08.041. PubMed PMID: WOS:000397295800044.
16. Therneau TM, Grambsch PM. Modeling survival data : extending the Cox model. New York: Springer; 2000. xiii, 350 p. p.
17. Therneau T. A Package for Survival Analysis in R. version 3.5-0 ed2023.
18. Halekoh U, Hojsgaard S, Yan J. The R Package geepack for Generalized Estimating Equations. J Stat Softw. 2006;15(2):1-11. doi: DOI 10.18637/jss.v015.i02. PubMed PMID: WOS:000235180600001.
19. Yan J, Fine J. Estimating equations for association structures. Stat Med. 2004;23(6):859-74. doi: 10.1002/sim.1650. PubMed PMID: WOS:000220103500001.
20. Datta D. blandr: a Bland-Altman Method Comparison package for R. . 0.5.1 ed2017.
21. Richardson DB, Ciampi A. Effects of exposure measurement error when an exposure variable is constrained by a lower limit. Am J Epidemiol. 2003;157(4):355-63. doi: 10.1093/aje/kwf217. PubMed PMID: WOS:000181057800011.
22. Tobin J. Estimation of Relationships for Limited Dependent Variables. Econometrica. 1958;26(1):24-36.
23. Mersmann O, Trautmann H, Steuer D, Bornkamp B. truncnorm: Truncated Normal Distribution. version 1.0-8 ed2018.
